# Changes in life expectancy and life span equality during the COVID-19 epidemic in Japan up to 2022

**DOI:** 10.1101/2024.04.18.24306049

**Authors:** Yuta Okada, Hiroshi Nishiura

**Affiliations:** Graduate School of Medicine, Kyoto University, Kyoto 6068503, Japan

**Keywords:** COVID-19, Life Expectancy, Life Tables, Cause of Death, Japan

## Abstract

**Objectives:** To evaluate the impact of COVID-19 on life expectancy in Japan through demographic analyses.

**Methods:** We evaluated the relationship between life expectancy gap from 2020-21 and 2021-22 and indicators of COVID-19 epidemic size at prefectural level. We also conducted age- and cause-specific decomposition of life expectancy change. Trends of life span equality from 2000-22 were also evaluated at the national level.

**Results:** Prefectural analysis between 2021-22 life expectancy change and annual per-population COVID-19 cases, person days in intensive care, reported COVID-19 deaths did not reveal significant correlations, which was contrary to our analysis from 2020-21. However, decomposition analysis revealed substantial life expectancy shortening attributable to the over-35-year-old population, and large increases in death causes such as cardiovascular or respiratory disorders along with COVID-19. Life span equality in Japan for the total population declined in 2020 but increased in 2021 and 2022 despite the shortening in life expectancy.

**Conclusions:** Discrepancy between life expectancy change and COVID-19 statistics in 2022 suggests the growing ascertainment bias of COVID-19. The increased contribution of cardiovascular disorders to life expectancy shortening is an alarming sign for the future. Life span equality in 2021 and 2022 is likely attributed to increased mortality among the elderly.

**Highlights:**

- Life expectancy change was not correlated with epidemic activity of COVID-19 in 2022
- Older people contributed to life expectancy shortening more than ever
- Cardiovascular disorders contributed substantially to life expectancy shortening
- Life span equality increased in 2021 and 2022 despite life expectancy shortening

## 1. Introduction

Since the start of the COVID-19 pandemic in Wuhan, China in November 2019, evidence of the pandemic’s impact on mortality has accumulated globally, with substantial geographical heterogeneity. (1–6) Global studies suggest that from January 1^st^, 2020 to December 31^st^, 2021, excess deaths worldwide were in the range of 14.9–15.9 million, with a large proportion attributed to India and the United States.(3,4) Published studies suggest that the global life expectancy change was −1.6 years from 2019 to 2021, when many countries showed bounce-backs from the shortening in 2020. However, other countries faced sustained shortening into 2021.(4–6)

It is now several years since the emergence of COVID-19, and the evaluation of the mortality impact of the condition has become more difficult for several reasons. One reason is changes in the official COVID-19 statistics, which are provided by public health agencies around the world and reflect epidemic activity. These are now less rigorous than in 2020, because most countries have gradually diminished their effort either to control the spread of COVID-19 or to maintain a meticulous surveillance system. Another reason is the change in the nature of deaths associated with COVID-19 since the introduction of vaccines against the disease in late 2020. The direct mortality impact of COVID-19 has been alleviated by these vaccines, but a substantial proportion of deaths are caused indirectly through complications such as cardiovascular disorders, or by limited access to healthcare services when the healthcare capacity or ambulance system were overwhelmed by the increased case load pressure of COVID-19. (5,7–15) The ongoing emergence of SARS-CoV-2 variants with a high capability of immune evasion and transmission may have worsened the health impact of COVID-19, but understanding the true burden has remained a challenging task.(16,17)

Direct approaches to estimating the mortality impact of COVID-19 are therefore challenging, including in Japan. There, the epidemic size of COVID-19 was greatest upon the emergence of SARS-CoV-2 Omicron (B.1.1.529) lineage variants. In line with other regions, Japan has been severely affected by COVID-19 in terms of excess mortality and life expectancy shortening. (1,3–5,14,18–21) The updated estimates by the National Institute of Population and Social Security Research suggest that life expectancy at birth has shortened for two consecutive years, from 84.58 years in 2021 to 84.10 in 2022 for the total population. However, it is not clear whether the cause-specific impact of this shortening has changed since 2021. It is also not clear how the contribution of cardiovascular, respiratory, and neoplastic disorders in 2021 have changed. (18) From a demographic perspective, the change in life span equality during and since the COVID-19 pandemic is also interesting. One measure of life span equality, or, evenness of life span, is the logarithm of the inverse of life table entropy. Global and historical demographic analysis suggests that the trends in life expectancy at birth and life span equality have been in line with each other. (22,23) However, this might not be the case when the age–mortality structure changes drastically. For example, during the COVID-19 epidemic in Japan, the mortality increase in 2021 contributed substantially to shorter life expectancy. (18)

To examine the demographic impact of the COVID-19 epidemic in 2022 in Japan, we investigated the relationship between reported COVID-19 burden at the prefectural level and life expectancy. We also decomposed the year-on-year life expectancy change from 2019–22 by age groups and major causes of death, and evaluated the lifetime loss by age and life span equality during the COVID-19 epidemic.

## 2. Material and methods

### 2.1. Epidemiological data

We used the data on deaths and exposure-to-risk populations available in the Japanese Mortality Database (JMD), which was available for the whole of Japan and by prefecture. (24) Death counts by cause of death and age group were obtained from the vital statistics published by the Ministry of Health, Labour and Welfare of Japan.(25) In line with a previous study, we categorized major causes of death using the International Statistical Classification of Diseases and Related Health Problems 10^th^ Revision (ICD- 10) into the top nine major cause categories (based on death counts by cause in 2022), and aggregated the remainder into a single group, to give a total of ten groups.(18,25) The epidemiological data for COVID-19 were retrieved from the open-access data provided by the Ministry of Health, Labour and Welfare. (26)

### 2.2. Calculation of period life table and Arriaga decomposition

For subsequent use for age- and cause-specific decomposition of life expectancy gaps, we re-calculated period life tables from 2000 to 2022 for the whole of Japan and for all prefectures as described before. (18) For the age- and cause-specific decomposition of life expectancy change, we used the Arriaga method for age- and cause-specific decomposition of life expectancy change. (18,27,28)

### 2.3. Life expectancy change and COVID-19 statistics at the prefectural level

Three COVID-19 statistics at prefectural level were used for this analysis: (i) annual number of COVID-19 cases, (ii) annual number of person-days in intensive care because of COVID-19, and (iii) annual number of documented deaths due to COVID-19. Using each of the COVID-19 indicators as an explanatory variable, we used linear regression analysis to predict the year-on-year life expectancy change as the dependent variable for 2020–21 and 2021–22.

### 2.4. Life span equality h

We used a measure of life span equality *h*, which was derived from life table entropy *H̅*. (22,29–31) We used a 1×1 year life table provided by JMD to calculate *h*(*t*) from 2000 to 2022 for total, male, and female populations, and evaluated the relationship between *h*(*t*) and life expectancy at birth, *e*_O_(*t*), for each of these populations. (24) As in Aburto et al. (22), we also calculated *w*(*x*, *t*)*W*_h_(*x*, *t*), the sensitivity of *h*(*t*) to the rate of mortality improvement, and its threshold age *a^H^* that satisfies *W* (*a^H^*, *t*) =0 for *t* = 2000, 2001, … , 2022. These results were compared with year-on-year mortality improvement, i.e., *r*(*x*, *t*) = log(*µ*(*a*, *t*))- log(*µ*(*a*, *t* + 1)), which is analogous to the rate of mortality improvement as described above.

### 2.5. Software

All analyses used R version 4.2.2.(32)

## Results

The summary of life expectancies at birth in Japan for the total, male, and female populations from 2019-2022 is shown in Supplementary Table 1. (Results based on abridged life tables that we re-calculated for use in Arriaga decomposition, which are almost identical to results provided by JMD) Life expectancy of the total population decreased by 0.49 years, from 84.59 to 84.10 from 2021-22. Though the change in the trend of life expectancy was already seen from 2020-21 by 0.15 years (from 84.74 to 84.59 years), the magnitude of shortening was greater in 2021-22. The shortening of life expectancy at birth for male and female populations also grew greater from 2021-22, estimated at 0.43 years (from 81.49 to 81.06 years) and 0.50 years (from 87.62 to 87.12 years), respectively.

Figure 1 shows life expectancy changes of the total population by prefecture in 2019–20, 2020-21, and 2021–2022. Following the drastic change from the overall increasing trend in 2019-20 to the sharply decreasing trend in 2020-21, all except one prefecture saw a decline in life expectancies from 2021-22. In 2022, the greatest decrease in life expectancy was seen in Iwate (1.00 years), and the only prefecture that enjoyed the increasing trend was Nagasaki (0.05 years). The prefecture level life expectancy changes of male and female population were in mostly in line with that of the total population. (For the details, see Supplementary Data)

**Figure 1.**
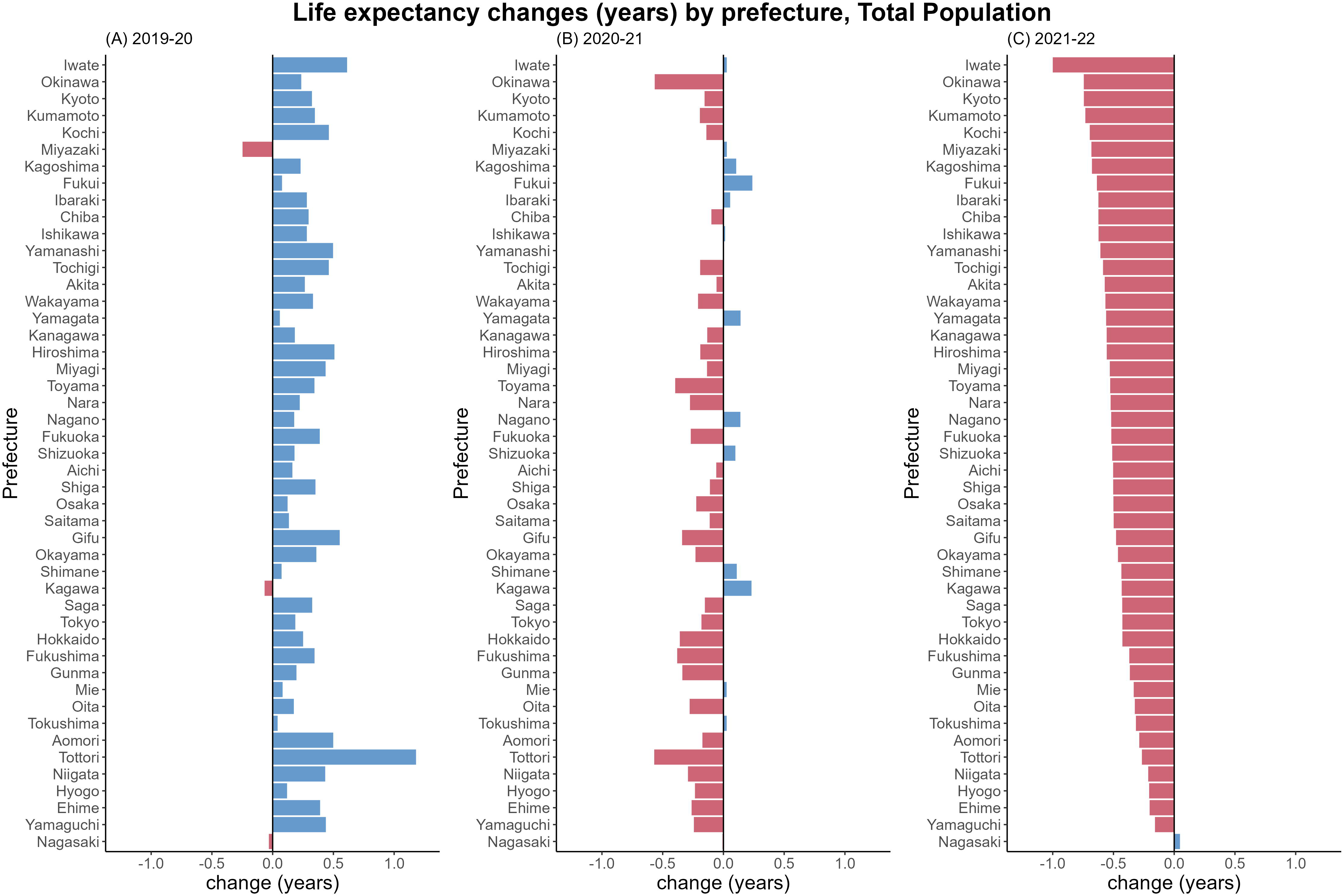
Life expectancy changes from 2019-20, 2020-21, and 2021-22 by prefecture. Changes from (A) 2019-20, (B) 2020-21, and (C) 2021-22 are shown. In each panel, bars in blue show positive changes, whereas red bars show negative changes. Order of prefectures are in an ascending order based on the life expectancy change of 2021-22.

Figure 2 presents the correlation between the reported COVID-19 burden and life expectancy changes at the prefectural level. Combined with the linear regression results as shown in Table 1, in 2021-22, the correlation between annual reported cases, person-day under intensive care, and death due to COVID-19 was not evident. This was contrary to the findings from 2020-21.

**Figure 2.**
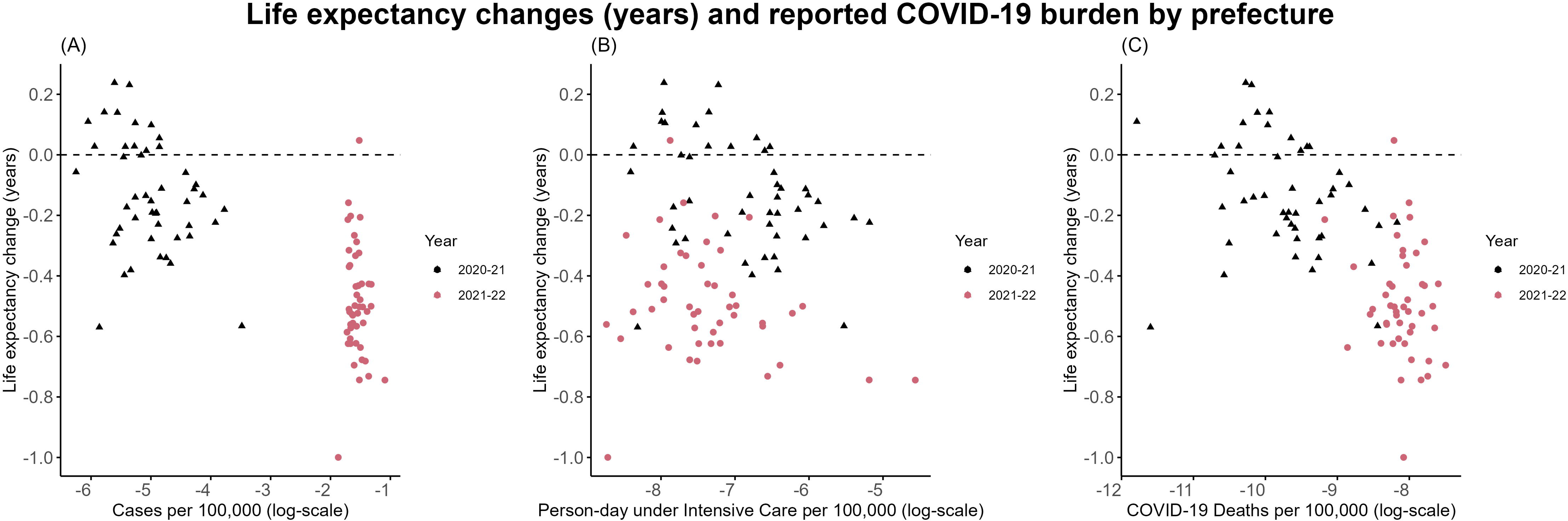
Correlation between life expectancy changes and COVID-19 burden based on official statistics. Correlation between life expectancy changes and the reported numbers of (A) annual COVID-19 cases, (B) person-day under intensive care due to COVID-19, and (C) deaths due to COVID-19 are shown. The variables on the x-axis are log-scaled in all panels. In each panel, individual prefectures are presented by black triangles representing 2020-21 data, or red dots representing 2021-22 data, respectively. The horizontal dashed line corresponds to “no year-on-year life expectancy change”.

**Table 1.** Life expectancy change and COVID-19 statistics: summary of linear regression analysis.

| COVID-19 data (log-scale) | Year | Coefficient (95% CI*) | Intercept (95% CI) |
| --- | --- | --- | --- |
| Cases | 2020-21 | -0.104 (-0.189, -0.018) | -0.655 (-1.086, -0.224) |
|  | 2021-22 | -0.104 (-0.496, 0.288) | -0.652 (-1.266, -0.037) |
| Person-days in intensive care | 2020-21 | -0.082 (-0.144, -0.021) | -0.704 (-1.132, -0.277) |
|  | 2021-22 | -0.053 (-0.117, 0.011) | -0.879 (-1.353, -0.405) |
| Death | 2020-21 | -0.067 (-0.137, -0.002) | -0.789 (-1.464, -0.114) |
|  | 2021-22 | -0.105 (-0.269, 0.060) | -1.338 (-2.671, -0.057) |

\*CI, confidence interval

Figure 3 shows the results of Arriaga decomposition of life expectancy change by age groups and major causes of death. (Aggregated summary by age groups or by causes of death are available as Supplementary Figures 1 and 2) As for the contribution by age, the negative contribution among the elderly population in 2020-21 was already seen. However, the negative contribution of the elderly population is more eminent in 2021-22 than in 2020-21. Also, the range of elder age with negative contribution has widened to younger quota in 2021-22 to as low as 30-34-year-olds.

**Figure 3.**
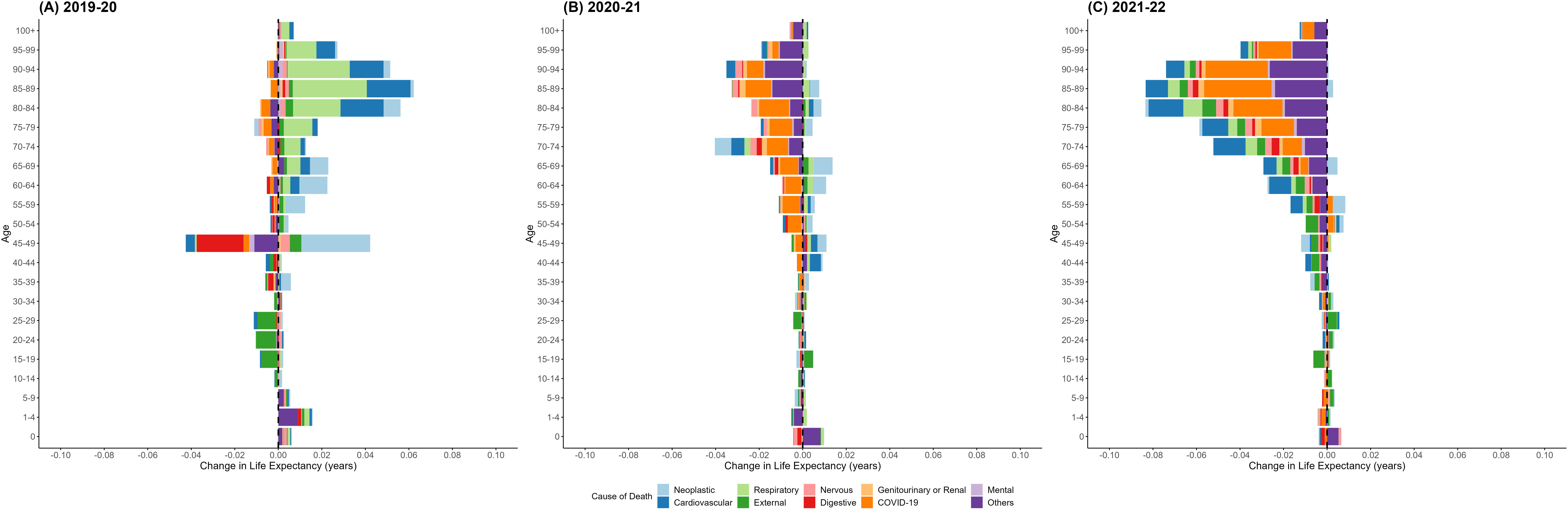
Arriaga decomposition of life expectancy change by major cause of death and age group, for the entire (total) population of Japan. Decomposed contribution by age for (A) 2019-20, (B) 2020-21, (C) 2021-22 are shown in each panel. Bar for each major cause is colored as shown in the panel below the plots. Bars representing major causes with positive contribution to life expectancy are stacked on the right-hand side, whereas those with negative contributions are stacked on the left-hand side.

As for contributions of major death causes by age groups which is also shown in Figure 3, negative contribution by COVID-19 among the elderly population has enlarged substantially in 2021-22, and the total contribution by all ages has grown from -0.095 years in 2020-21 to -0.131 years in 2021-22. In addition to COVID-19, the negative contribution of cardiovascular causes also grew much in 2021-22 especially among age groups older than 50 years old. The total contribution of cardiovascular death was -0.091 years in 2022 and has decreased consistently and substantially compared with +0.073 years in 2020 and -0.003 years in 2021. The negative contribution of “other” causes (the remainder of death causes that do not belong to the top 9 major death cause categories) also increased substantially in 2021-22 among elderly population older than 50 years old, with a total of -0.139 years across all age groups. The decrease in the contribution of respiratory and neoplastic disorders as well as that from other causes from 2020-21 to 2021-22 is also clearly observed. (See Supplementary Data for detailed results). Results from the decomposition analysis for the male and female populations were similar to that of the total population. (Supplementary Data and Supplementary Figures 3 and 4)

The values of *h*(*t*) as an indicator of life span equality for the total population from 2000 to 2022 are shown in Figure 4. As shown in panel (A), *h* has mostly increased monotonously up to 2019, with the exception of 2011 when an exceptional number of casualties occurred due to the earthquake and tsunami that hit eastern Japan. That increasing trend was halted in 2020 when the COVID-19 pandemic started, but has resumed to increase since 2021. The values of *h*(*t*) for female and male populations also showed very similar patterns to those for the total population. (Supplementary Figures 5 and 6) Panel (B) in Figure 4 shows the relationship between *h*(*t*) and life expectancy at birth from 2000 to 2022. A decrease in *h*(*t*) was seen in 2020 for the first time since 2011, and it was followed by an increase in 2021 and 2022 decreased despite the shortening of life expectancy at birth.

**Figure 4.**
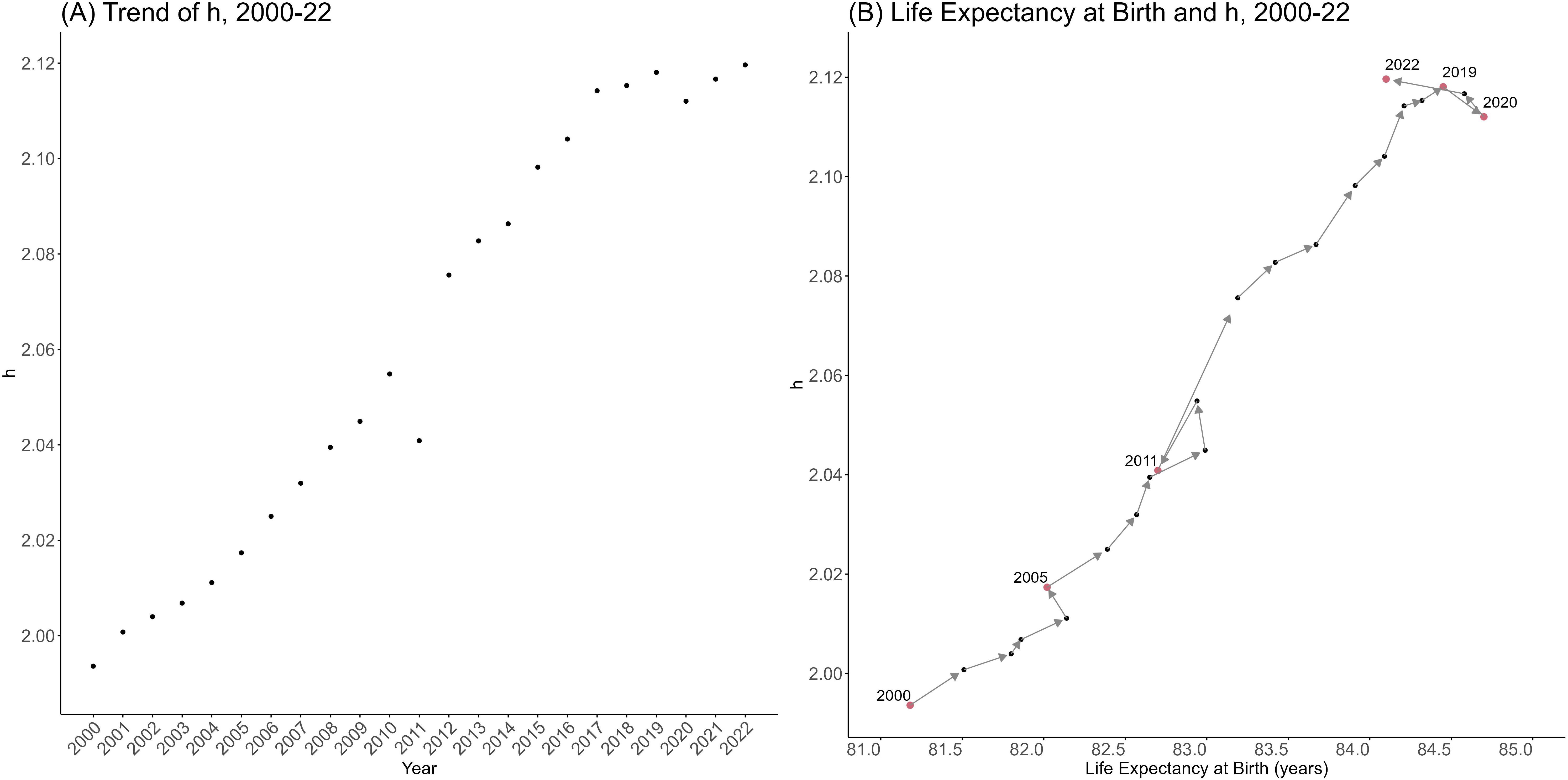
The trend of life span equality from 2000 to 2022, for the entire (total) population of Japan. Panel (A) shows the dynamics of life span equality along time from 2000 to 2022. Panel (B) shows the same dynamics in relation to life expectancy for the same period, where the years corresponding to the red dots are noted within the figure.

To see how the overall dynamics of *h*(*t*) from 2020 to 2022 can be explained by mortality improvements by age for this period (in relation to *a^H^*), we calculated the curves of *w*(*x*, *t*)*W_h_*(*x*, *t*) across ages for 2021 and 2022, and also evaluated the year- on-year mortality improvement *r*(*x*, *t*) from 2020 to 2022 for the total population. (Supplementary Figure 6) The curves of *w*(*x*, *t*)*W_h_*(*x*, *t*) for 2021 and 2022 were very similar, although a slight shift toward the younger ages can be observed in the negative part of the curve in the elderly population. As for *r*(*x*, *t*), *r*(*x*, 2020) above *x*= *a^H^* lied in the positive range, whereas *r*(*x*, 2021) and *r*(*x*, 2022) mostly lied in the negative range for the age range. For ages younger than *x*= *a^H^*, the signs of *r*(*x*, 2020), *r*(*x*, 2021), and *r*(*x*, 2022) were inconsistent across different ages, suggesting that increased mortality among ages older than *a^H^* have clearly contributed to the increase of *h*(*t*) in 2021 and 2022. (Also see Supplementary Figures 7 and 8 for results on female and male populations)

## Discussion

Our study showed the pattern of deaths in Japan during the COVID-19 epidemic (up to 2022) through demographic information. The main finding was the growing impact of the older population and cardiovascular deaths on the shortening of life expectancy, which was considerable from 2021 to 2022. The lack of significant correlations between life expectancy change and epidemiological indicators of the COVID-19 burden from 2022 is also a concern. This finding may be linked to the low detection of COVID-19 cases and associated deaths, which is supported by our results about the age- and cause-specific contributions to life expectancy change. The increasing trend in life span equality despite the life expectancy shortening may also be related to the substantial increase in mortality among the older population.

There were two key findings from our study. The first was that all age groups over 30 years old contributed to the shortening of life expectancy in 2022, as shown in Figure 3 and Supplementary Figure 1. However, compared with the overall shortening attributed to age groups over 50 in 2021, the negative impact was more diffuse across ages. This finding is similar to what was observed in 2020–21 in countries in Eastern Europe, though the underlying situations in these countries, such as types of circulating SARS-CoV-2 variants, vaccine coverage, and healthcare situations, would have been quite different from that in Japan from 2021–22. (5) In Japan, the population-wide vaccine coverage of the second dose of mRNA vaccines (BNT162b2 [Pfizer/BioNTech] and mRNA-1273 [Moderna] vaccines) was around 80% by the end of 2021, and the coverage of the third dose also increased from around 15% at the end of 2021 to 68% by the end of 2022. (33) Despite this high vaccination rate, we found substantial mortality caused by COVID-19 in Japan among wider age groups in 2022. This was not fully captured by COVID-19 statistics, as seen in our prefectural analyses (Figure 2 and Table 1).

Another key finding was the substantial growth in the negative contribution of cardiovascular disorders to life expectancy shortening, especially among populations over 50 years old (Figure 3 and Supplementary Figure 2). This was not surprising, because published studies have shown an elevated risk of cardiovascular diseases associated with COVID-19. (12–14,34) However, to our knowledge, our study is the first to have quantified the magnitude of life expectancy shortening in Japan caused by cardiovascular deaths in 2022. The negative change in contributions by respiratory causes from 2021 to 2022 and the consistently negative trend in contributions by neoplastic disorders since 2020 are also of note. In addition to COVID-19-associated conditions, these findings may be attributable to an array of factors including changes in hospital attendance. (18) There is a gap between these findings and the global and regional cause-specific contributions to life expectancy change from 2019–21. (35) Further update on this issue is warranted to evaluate changes in life expectancy change by causes of death. The increase in the contribution of remaining causes of death is mostly explained by the increase in deaths due to senility, which increased by around 20,000 in 2021 and 27,000 in 2022. (25)

The changes in life span equality during the COVID-19 pandemic were also of note. Our result highlights the undesirable increase in life span equality despite the shortening of life expectancy at birth. This was in line with the substantial negative contribution by the older population to life expectancy changes in the same period, highlighted by the Arriaga decomposition results. These findings add to demographic case studies on the historical relationship between life expectancy and life span equality. (22,23)

Our study had some limitations. First, we could not examine the relationship between COVID-19 and other causes of death in detail at the prefectural level, because data on prefectural death count stratified by age and cause of death are not openly accessible. Detailed analysis of prefectural data would have provided insights on geographic heterogeneity, and we hope to explore this in future. Second, we ignored geographic and temporal variation in the ascertainment bias for COVID-19 statistics. We sufficiently met our key focus to be confident about the true mortality burden of COVID-19, but these factors could have biased our analysis of the relationship between prefectural COVID-19 statistics and life expectancy change. Third, we did not consider the fluctuation in the coverage of death registrations in Japan from 2019–22. However, it is unlikely that we missed a large proportion of deaths that would substantially affect our results, because the completeness of death registration is reported to be 90–99% in Japan. (36)

In conclusion, our demographic analysis showed the impact of the COVID-19 epidemic up to 2022, when the epidemic grew substantially larger. The demographic burden of the pandemic increased more in 2022 than in 2021 or before, but the COVID- 19 burden reported by epidemiological surveillance failed to capture this trend. This is probably due to both the shrinking coverage of epidemiological surveillance and the growing impact of COVID-19-associated deaths caused by complications such as cardiovascular disorders. We also showed an undesirable increase in life span equality due to disproportionately higher mortality among older people. Our study therefore provides valuable insights into the mortality impact of the COVID-19 epidemic in Japan, which can now only be captured by indirect measures such as demographic analysis in the absence of meticulous epidemiological surveillance.

## Data Availability

All data produced in the present work are contained in supplementary materials

https://www.mhlw.go.jp/stf/seisakunitsuite/newpage_00023.html

https://www.fdma.go.jp/disaster/coronavirus/post-1.html

https://www.mhlw.go.jp/stf/seisakunitsuite/bunya/0000121431_00432.html

https://www.ipss.go.jp/p-toukei/JMD/index-en.asp

https://www.e-stat.go.jp/en/stat-search/files?page=1&layout=datalist&toukei=00450011&tstat=000001028897&cycle=7&tclass1=000001053058&tclass2=000001053061&tclass3=000001053065&cycle_facet=tclass1&tclass4val=0

https://covid19.mhlw.go.jp/en/

https://www.kantei.go.jp/jp/headline/kansensho/vaccine.html

https://unstats.un.org/unsd/demographic-social/crvs/documents/2023-completeness.xlsx

## Acknowledgment

We thank Melissa Leffler, MBA, of Edanz (https://jp.edanz.com/ac) for editing a draft of this manuscript.

## Ethical approval statement

Ethical approval was not required because all data used in the present study did not include any personally identifiable information.

## Data availability

We used openly accessible COVID-19 statistics from the website of the Ministry of Health, Labour or Welfare, and life tables and related statistics from the website of National Institute of Population and Social Security Research. The supplementary files include the datasets used in this study, and also the results of our numerical analyses. None of the data used in the present study contained personally identifiable information.

---

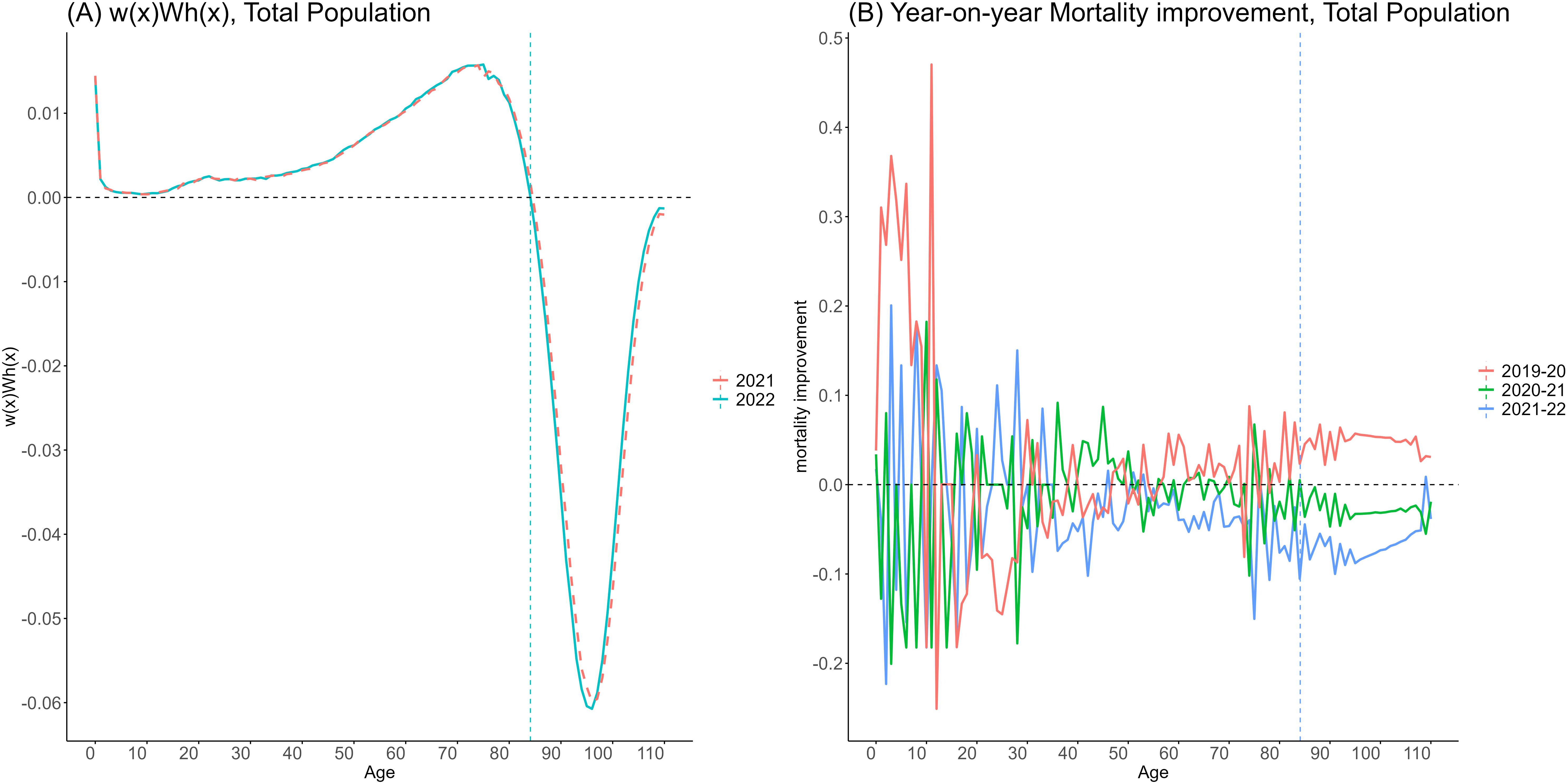

