## supplementary material for "Changes in life expectancy and life span equality during the COVID-19 epidemic in Japan up to 2022"

**Supplementary Figures**

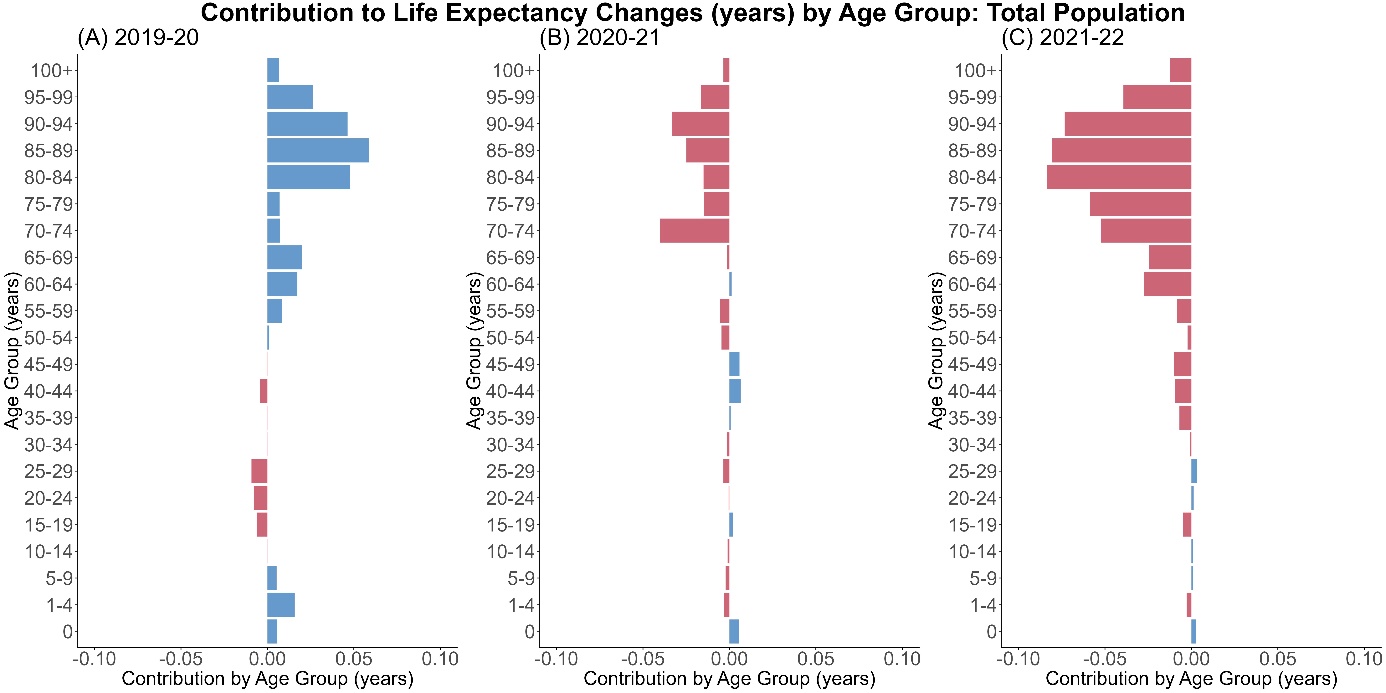

**Supplementary Figure 1. Arriaga decomposition of life expectancy change by age group for the total population of Japan.**

Decomposed contribution by age for (A) 2019–20, (B) 2020–21, (C) 2021–22 are shown in each panel. Blue bars show positive contributions, and red bars negative contributions.

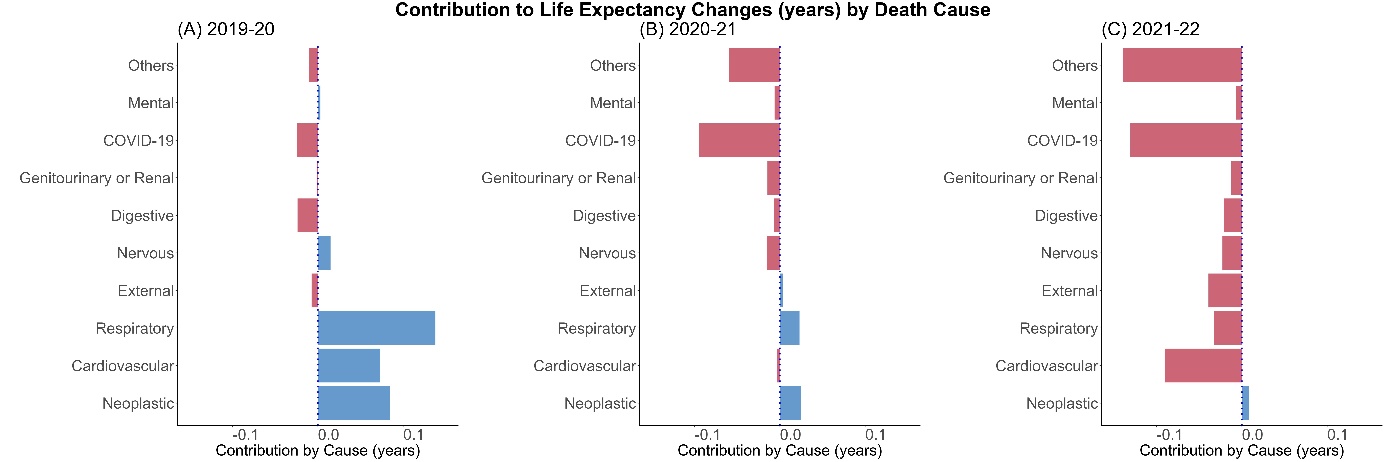

**Supplementary Figure 2. Arriaga decomposition of life expectancy change by major causes of death of Japan.**

Decomposed contribution by age for (A) 2019–20, (B) 2020–21, (C) 2021–22 are shown in each panel. As in Supplementary Figure 1, blue bars show positive contributions, and red bars negative contributions.

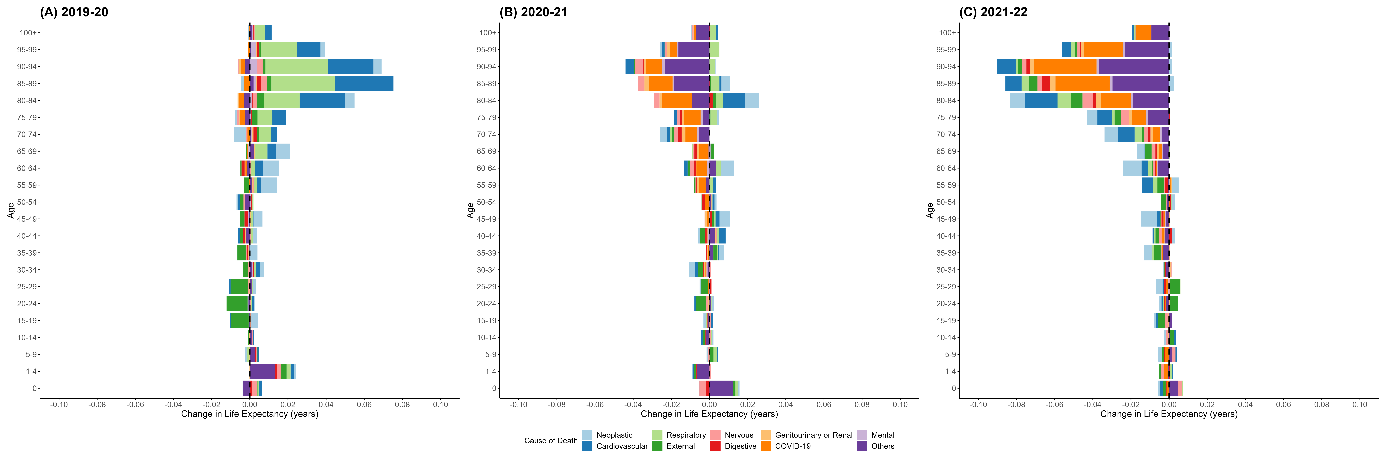

**Supplementary Figure 3. Arriaga decomposition of life expectancy change by major cause of death and age group, for the female population of Japan.**

Decomposed contribution by age for (A) 2019–20, (B) 2020–21, (C) 2021–22 are shown in each panel. The key for the colors of the bars is shown in the panel below the plots. Bars for major causes with positive contributions to life expectancy are stacked on the right-hand side, and those with negative contributions are on the left-hand side.

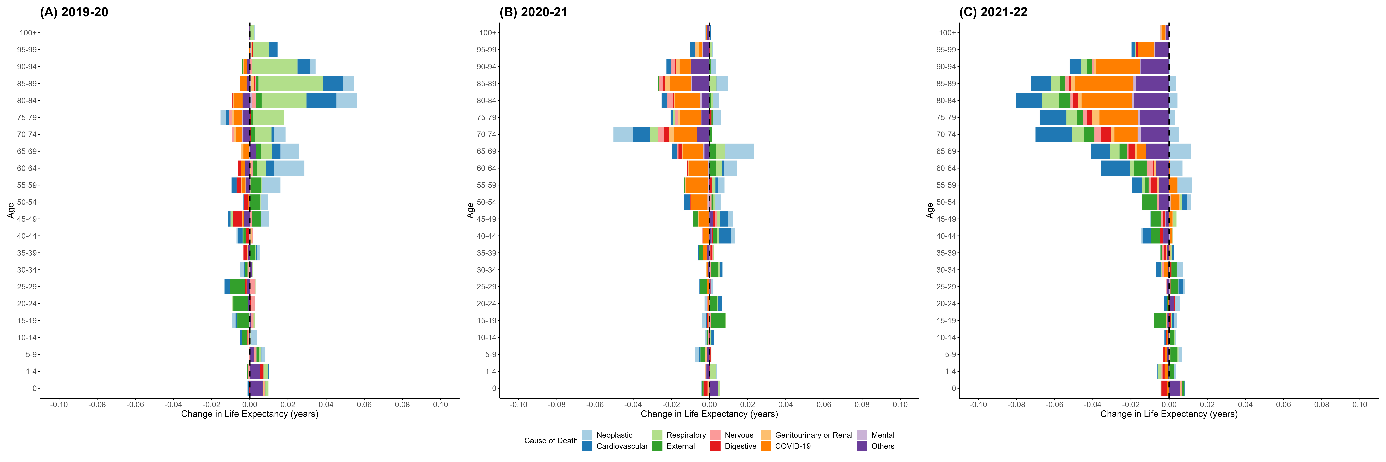

**Supplementary Figure 4. Arriaga decomposition of life expectancy change by major cause of death and age group, for the male population of Japan.**

Decomposed contribution by age for (A) 2019–20, (B) 2020–21, (C) 2021–22 are shown in each panel. The key for the bar colors are shown in the panel below the plots. Bars representing major causes with positive contributions to life expectancy are stacked on the right-hand side, and those with negative contributions are on the left-hand side.

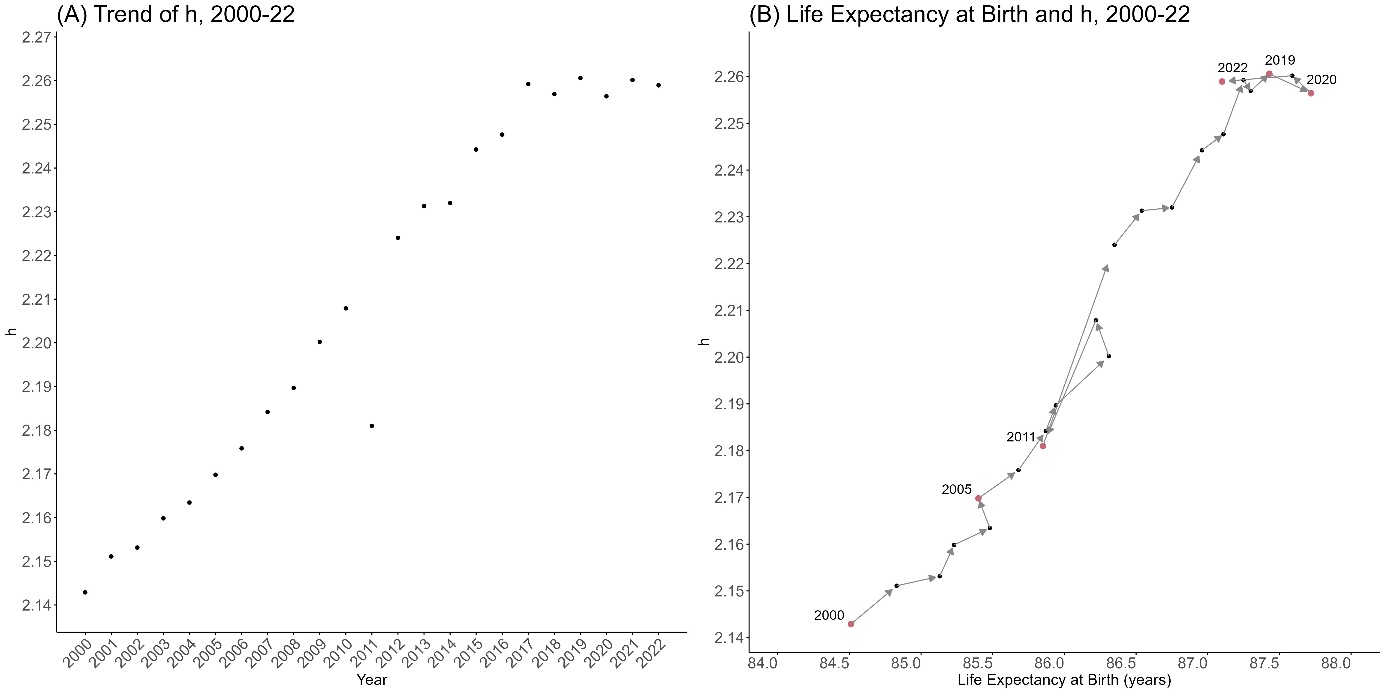

**Supplementary Figure 5. The trend in life span equality from 2000 to 2022, female population of Japan.**

Panel (A) shows the dynamics of life span equality by time from 2000 to 2022. Panel (B) shows the same dynamics in relation to life expectancy for the same period. The years corresponding to the red dots are noted within the figure.

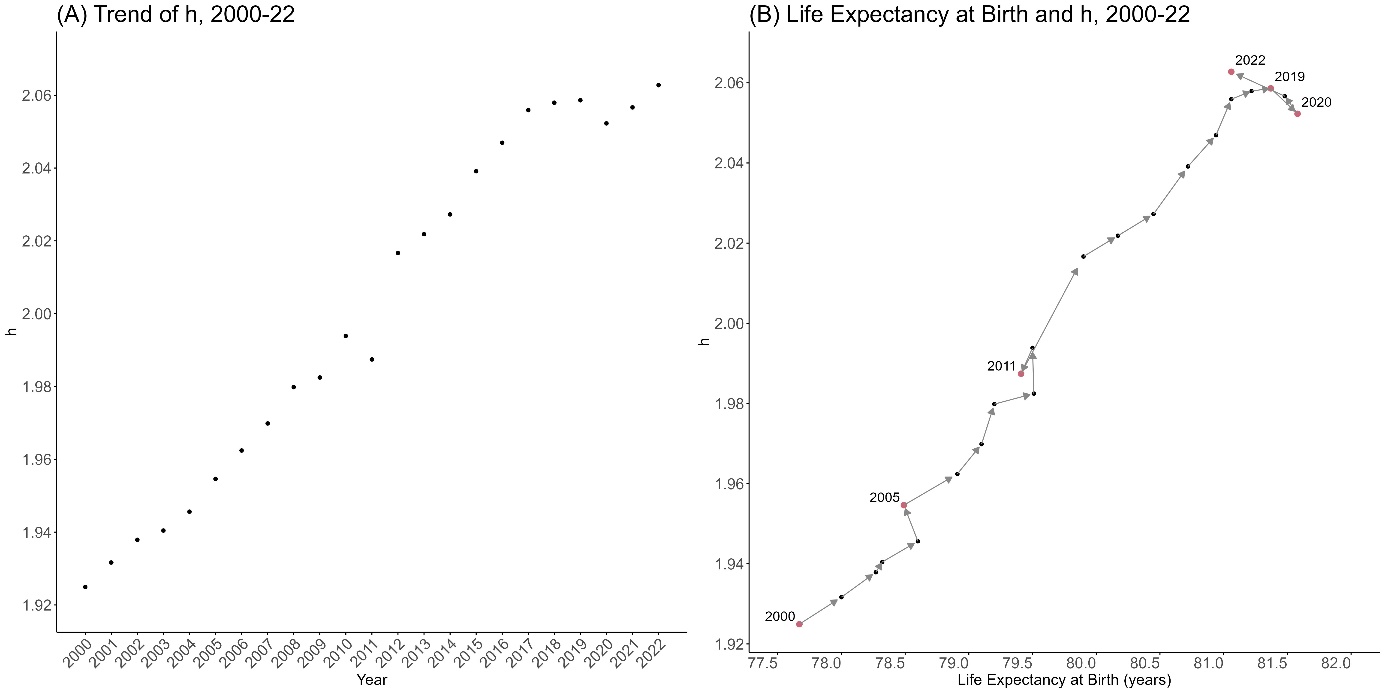

**Supplementary Figure 6. The trend in life span equality from 2000 to 2022, male population of Japan.**

Panel (A) shows the dynamics of life span equality by time from 2000 to 2022. Panel (B) shows the same dynamics in relation to life expectancy for the same period, and the years corresponding to the red dots are noted within the figure.

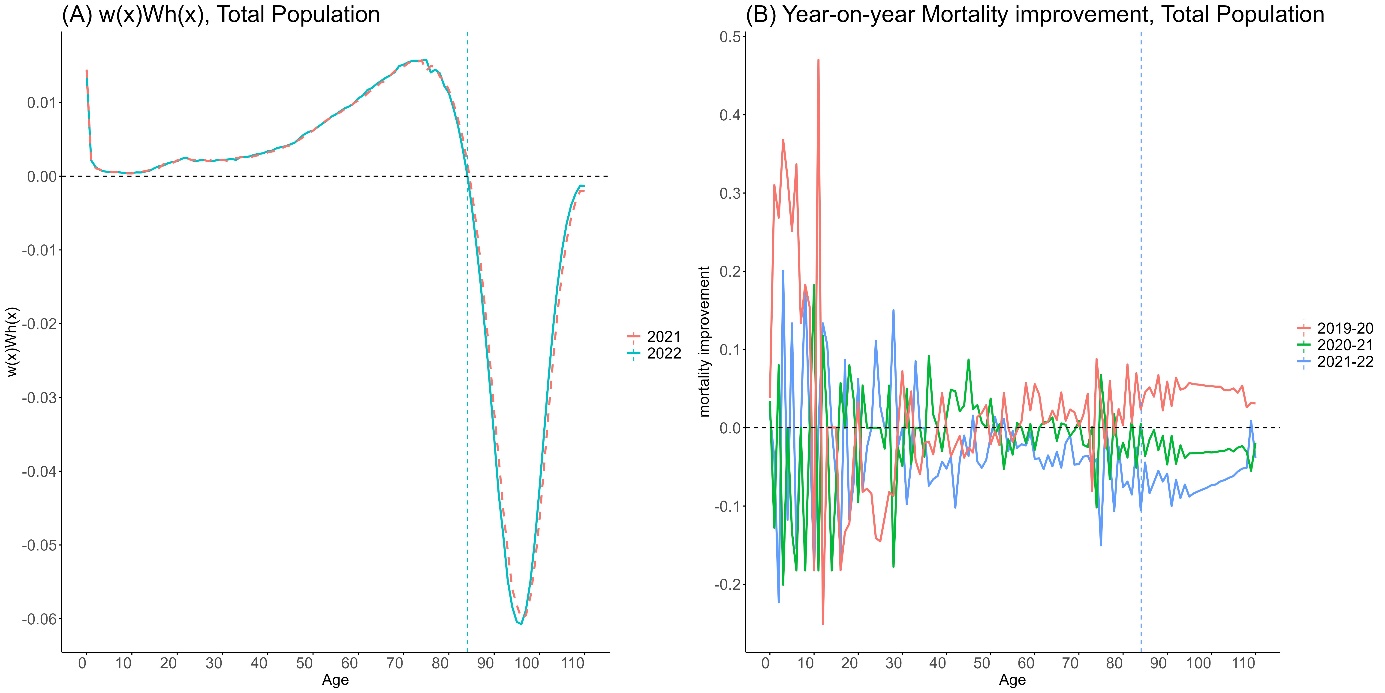

**Supplementary Figure 7. Weight of change in life span equality and mortality improvement by age in Japan, total population.**

Panel (A) shows the weight $w\left( x,t \right)W_{h}\left( x,t \right)$ for $t=2022$ (solid blue line) and $t=2021$ (dashed red line). Panel (B) describes the year-on-year mortality improvement $r\left( x, t \right)=\log\left( \mu\left( a, t \right) \right)-\log\left( \mu\left( a, t+1 \right) \right)$ for $t=2020$ (red), $t=2021$ (green), and $t=2022$ (blue). Vertical dashed lines in both panels represent the threshold age $a^{H}=84.10$ for 2022.

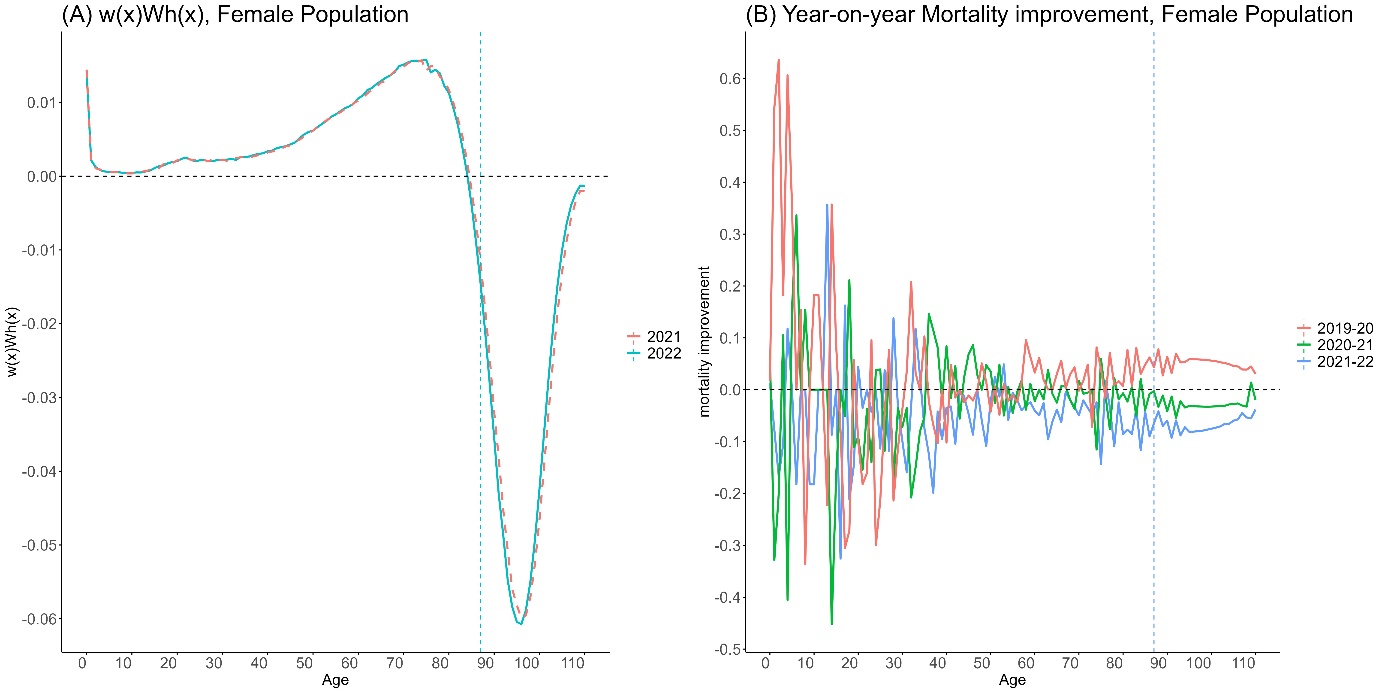

**Supplementary Figure 8. Weight of change in life span equality and mortality improvement by age in Japan, female population.**

Panel (A) shows the weight $w\left( x,t \right)W_{h}\left( x,t \right)$ for $t=2022$ (solid blue line) and $t=2021$ (dashed red line). Panel (B) describes the year-on-year mortality improvement $r\left( x, t \right)=\log\left( \mu\left( a, t \right) \right)-\log\left( \mu\left( a, t+1 \right) \right)$ for $t=2020$ (red), $t=2021$ (green), and $t=2022$ (blue). Vertical dashed lines in both panels represent the threshold age $a^{H}=84.10$ for 2022.

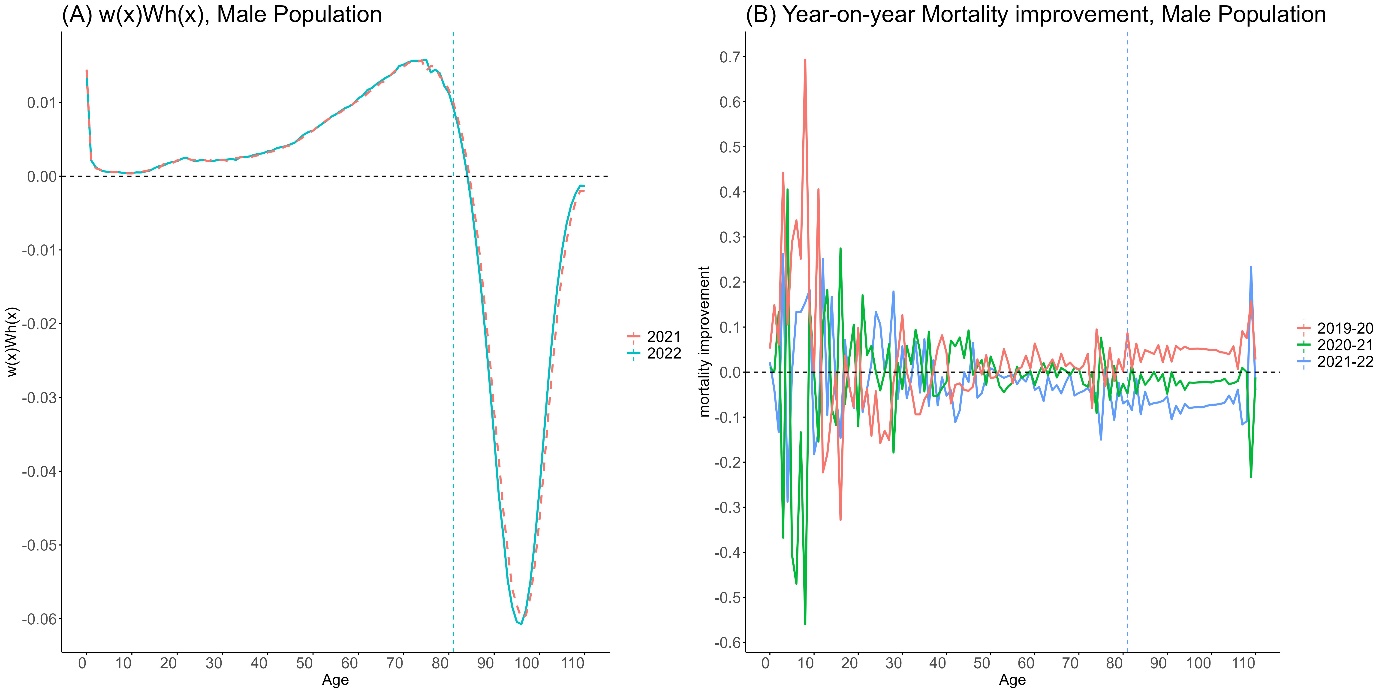

**Supplementary Figure 9. Weight of change in life span equality and mortality improvement by age, male population.**

Panel (A) shows the weight $w\left( x,t \right)W_{h}\left( x,t \right)$ for $t=2022$ (solid blue line) and $t=2021$ (dashed red line). Panel (B) describes the year-on-year mortality improvement $r\left( x, t \right)=\log\left( \mu\left( a, t \right) \right)-\log\left( \mu\left( a, t+1 \right) \right)$ for $t=2020$ (red), $t=2021$ (green), and $t=2022$ (blue). Vertical dashed lines in both panels represent the threshold age $a^{H}=84.10$ for 2022.

**Supplementary Table**

**Supplementary Table 1. Life expectancy of total, male, and female populations in Japan, 2019–22.**

| Population | Total | Male | Female |
| --- | --- | --- | --- |
| 2019 | 84.49 | 81.41 | 87.48 |
| 2020 | 84.74 | 81.61 | 87.77 |
| 2021 | 84.59 | 81.49 | 87.62 |
| 2022 | 84.10 | 81.06 | 87.12 |
